# Baseline Cardiometabolic Profiles and SARS-CoV-2 Infection in the UK Biobank

**DOI:** 10.1101/2020.07.25.20161091

**Authors:** Ryan J. Scalsky, Yi-Ju Chen, Karan Desai, Jeffery R. O’Connell, James A. Perry, Charles C. Hong

## Abstract

**Background:** SARS-CoV-2 is a rapidly spreading coronavirus with a high incidence of severe upper respiratory infection that first presented in Wuhan, China in December 2019. Many factors have been identified as risk factors for SARS-CoV-2, with much attention being paid to body mass index (BMI). Little investigation has been done to investigate dysregulation of lipid profiles and diabetes, which are often comorbid in high BMI patients.

**Objective:** This study seeks to describe the impact of BMI, HDL, LDL, ApoA, ApoB, triglycerides, hemoglobin A1c (HbA1c), diabetes, alcohol and red wine intake on the odds of testing positive for SARS-CoV-2 in UK Biobank (UKB) study participants.

**Methods:** We examined the effect of BMI, lipid profiles, diabetes and alcohol intake on the odds of testing positive for SARS-Cov-2 among 9,005 UKB participants tested for SARS-CoV-2 from March 16 through July 14, 2020. Odds ratios and 95% confidence intervals were computed using logistic regression adjusted for age, sex and ancestry.

**Results:** Higher BMI, Type II diabetes and HbA1c were associated with increased SARS-CoV-2 odds (p < 0.05) while HDL and ApoA were associated with decreased odds (p < 0.001). Though the effect of BMI, Type II diabetes and HbA1c were eliminated when HDL was controlled, the effect of HDL remained significant when BMI was controlled for. Additionally, red wine intake was associated with reduced odds of testing positive for SARS-CoV-2 (p < 0.05). LDL, ApoB and triglyceride levels were not found to be significantly associated with increased odds.

**Conclusion:** Elevated HDL and ApoA levels and alcohol intake, specifically red wine intake, were associated with reduced odds of testing positive for SARS-CoV-2, while higher BMI, type II diabetes and HbA1c were associated with increased odds. The effects of alcohol, BMI, type II diabetes and HbA1c levels were no longer significant after controlling for HDL, suggesting that these effects may be mediated in part through regulation of HDL levels. In summary, our study corroborates the emerging picture that high HDL levels may confer protection against SARS-CoV-2.

**Highlights:**

- Higher baseline HDL levels were associated with reduced odds of testing positive for SARS-CoV-2.
- BMI, Type II diabetes and hemoglobin A1C levels were associated with elevated odds of testing positive for SARS-CoV-2, but this effect was abrogated when controlling for HDL.
- Red wine intake was associated with reduced odds of testing positive for SARS-CoV-2, although this effect may in part be moderated by HDL.
- Baseline LDL and Triglyceride levels were not associated with increased odds of testing positive for SARS-CoV-2.

## Introduction

Since early December 2019, when the first cases of the severe acute respiratory syndrome coronavirus 2 (SARS-CoV-2 or Covid-19) were identified in Wuhan, China, nearly 13 million individuals have tested positive for the virus (1). Researchers have rapidly attempted to define the clinical characteristics associated with increased risk of becoming infected with SARS-CoV- 2 to improve our understanding and clinical management of this pandemic. Early data from across the globe have identified pre-existing cardiovascular disease and obesity as risk factors associated with acquiring SARS-CoV-2 (2-8). Curiously, early data from China also found that hypolipidemia and declining HDL at the time of acute COVID19 illness was associated with disease severity (9, 10). There remains limited research on how an individual’s baseline cardiometabolic profile, specifically lipid levels, affect one’s risk for contracting the virus. This has received particular attention as the known viral entry mechanism for SARS-CoV-1, a closely related virus responsible for the 2003 SARS outbreak in China, has been shown, in preliminary in-vitro studies, to be cholesterol-dependent (11). In this paper we analyze the association of a positive SARS-CoV-2 test with an individual’s lipid profile in the UK Biobank resource.

## Methods

The UK Biobank resource began releasing SARS-CoV-2 test results in April 2020 to approved researchers. Full details on these test results are available online (12). Using test results released on July 14, 2020, we classified subjects testing positive for SARS-CoV-2 as cases. If multiple tests were performed, we classified a subject as a case if any test gave a positive result, based on the rationale that false positives are less likely than false negatives. Those with only negative test results were classified as controls. Tests were initially conducted in hospital settings in individuals who presented with respiratory symptoms. From April 27^th^ onward, testing was expanded to include community clinics and all non-elective patients admitted overnight, including those who were asymptomatic. 70.1% of the 9,005 subjects (cases and controls) were inpatient when the sample was taken, 74.1% of 7,497 controls were inpatient when the sample was taken and 50.3% of 1,508 cases were inpatient when the sample was taken. The vast majority of samples for testing were obtained by nose/throat swabs and samples were analyzed for SARS-CoV-2 RNA via PCR.

The association analysis was performed with Plink2 (13) using logistic regression. The binary outcome variable of “SARS-CoV-2 test status” (cases tested positive, controls tested negative) was run against a series of independent variables which included continuous, categorical, and binary ICD10 data supplied by the UK Biobank. The data for continuous and categorical traits were collected when subjects were enrolled into the UK Biobank (2006-2010). Serum was collected for analysis of LDL, HDL, ApoA, ApoB and triglycerides. LDL was analyzed by enzymatic selective protection, HDL was analyzed by enzyme immunoinhibition, ApoA and ApoB were analyzed by immunoturbidimetric analysis and triglycerides were measured by GPO-POD. All analyses were completed using the AU5800 by Beckman Coulter. Height and weight were measured, and BMI was computed from these values. Impedance BMI was measured by bioelectrical impedance using the Tanita BC418MA body composition analyzer. Additional details on collection and analysis of biomarkers (e.g. LDL, HDL, ApoA) are available from the UK Biobank website (14). ICD10 diagnostic codes are current for all subjects through October 2019. We also grouped the ICD10 code data into Phecodes in order to increase statistical power (15). The analysis included covariates of sex, age, and principal components (PCs) 1 through 4 to adjust for ancestry. Principal component analysis, a standard technique used in statistical genetics, generates a dataset of PCs (typically 10) that can be used as covariates to correct for population stratification (i.e. differences in ancestry) (16). PCs provided by the UK Biobank, which were computed from the cohort’s genotypes, were used. Our preliminary analysis showed that only the first 4 PCs were significant at p < 0.05 and thus we included only PC1-4 as covariates.

Our analysis yielded odds ratios (OR) and 95% confidence intervals (CI) for each trait tested against the “SARS-CoV-2 test status”. An OR greater than 1.0 indicates increased odds of a SARS-CoV-2 positive test compared to the controls. An OR less than 1.0 indicates decreased odds. For continuous phenotypes, the OR indicates the increased odds (for OR > 1.0) or decreased odds (for OR < 1.0) per standard deviation increase in the continuous phenotype.

## Results

### Demographics

This dataset includes 1,508 cases and 7,497 controls for a total of 9,005 subjects. Prior to the association analysis we compared the cases and controls for differences in sex, ancestry and age. Significant differences were found between the sex (p-value = 1.3×10^−3^; Table 1), ancestry (p-value = 1.1×10^−15^; Table 1) and age (p-value = 3.4×10^−8^; Table 1) of cases and controls.

**Table 1.**
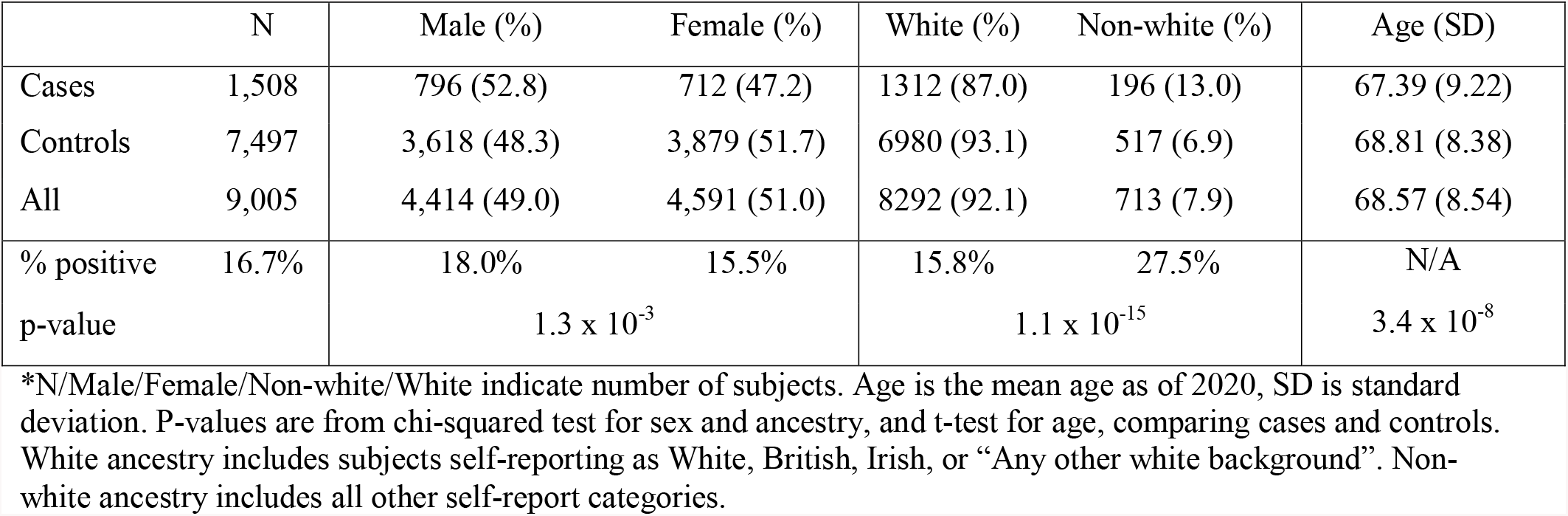
Demographics - Sex, Ancestry and Age.

### BMI

Body mass index (BMI) has been shown to increase SARS-CoV-2 risk across many populations (3). Interestingly, we found that BMI was associated with an increased odds of SARS-CoV-2 positive testing (OR = 1.12, 95% CI = 1.06 – 1.18, p-value = 6.14×10^−5^; Table 2) but when HDL was controlled for this effect was no longer significant (OR = 1.06, 95% CI = 0.995 – 1.13, p-value = 0.071; Table 2). These findings were consistent when BMI was measured by electrical impedance (OR = 1.12, 95% CI = 1.06 – 1.18, p-value = 8.54×10^−5^; Table 2) and the significance was also lost when HDL was controlled for (OR = 1.06, 95% CI = 0.994 – 1.13, p-value = 0.077; Table 2).

**Table 2.** Effect of Body Mass Index.

| Trait | Covariates | N | Cases/Controls | Mean (S.D.) | Odds Ratio | 95% Confidence Interval | p-value |
| --- | --- | --- | --- | --- | --- | --- | --- |
| BMI | Age, Sex, 4pc | 8,939 | 1,496/7,443 | 28.29 (5.27) | 1.12 | 1.06 – 1.18 | **$6.14 \times 10^{-5}$** |
| BMI | Age, Sex, 4pc, HDL | 7,764 | 1,280/6,484 | 28.31 (5.26) | 1.06 | 0.995 – 1.13 | 0.071 |
| Impedance BMI | Age, Sex, 4pc | 8,739 | 1,463/7,276 | 28.29 (5.26) | 1.12 | 1.06 – 1.18 | **$8.54 \times 10^{-5}$** |
| Impedance BMI | Age, Sex, 4pc, HDL | 7,590 | 1,252/6,338 | 28.30 (5.25) | 1.06 | 0.994 – 1.13 | 0.077 |

\*Body Mass Index (BMI) measured by height and weight in units of $\text{Kg/m}^2$ , Impedance BMI measured in increments of 0.1 in units of $\text{Kg/m}^2$

### HDL/ApoA

After controlling for age, sex and 4pc, we found that plasma HDL levels were associated with a reduced odds of testing positive for SARS-CoV-2 (OR = 0.845, 95% CI = 0.788 – 0.907, p-value = 2.45×10^−6^; Table 3), this effect was maintained even when controlling for BMI (OR = 0.863, 95% CI = 0.801 – 0.93, p-value = 1.17×10^−4^; Table 3). Moreover, we found that plasma levels of Apolipoprotein A (ApoA), the major protein component of HDL particles in plasma, were also associated with a reduced odds of testing positive for SARS-CoV-2 (OR = 0.849, 95% CI = 0.793 – 0.910, p-value = 2.90×10^−6^; Table 3). The effect of ApoA also remained significant when controlling for BMI (OR = 0.865, 95% CI = 0.806 – 0.929, p-value = 6.97×10^−5^; Table 3). Consistent with high collinearity between HDL and ApoA, the effect of either is negligible when the opposite is controlled for suggesting that both describe the same effect (Table 3).

**Table 3.** Effect of HDL and ApoA.

| Trait | Covariates | N | Cases/Controls | Mean (S.D.) | Odds Ratio | 95% Confidence Interval | p-value |
| --- | --- | --- | --- | --- | --- | --- | --- |
| HDL | Age, Sex, 4pc | 7,821 | 1,291/6,530 | 1.40 (0.38) | 0.845 | 0.788 – 0.907 | **$2.45 \times 10^{-6}$** |
|  | Age, Sex, 4pc, ApoA | 7,777 | 1,286/6,491 | 1.39 (0.37) | 0.944 | 0.804 – 1.11 | 0.480 |
|  | Age, Sex, 4pc, BMI | 7,764 | 1,280/6,484 | 1.40 (0.38) | 0.863 | 0.801 – 0.93 | **$1.17 \times 10^{-4}$** |
| ApoA | Age, Sex, 4pc | 7,783 | 1,288/6,495 | 1.51 (0.27) | 0.849 | 0.793 – 0.910 | **$2.90 \times 10^{-6}$** |
|  | Age, Sex, 4pc, HDL | 7,777 | 1,286/6,491 | 1.51 (0.27) | 0.894 | 0.762 – 1.048 | 0.167 |
|  | Age, Sex, 4pc, BMI | 7,727 | 1,277/6,450 | 1.51 (0.27) | 0.865 | 0.806 – 0.929 | **$6.97 \times 10^{-5}$** |

\*High density lipoprotein (HDL) measured in mmol/L, Apolipoprotein A (ApoA) measured in g/L

### Hyperlipidemia/LDL/ApoB/Triglycerides

The diagnosis of hyperlipidemia (ICD10 codes E78.4 and E78.5) was modestly associated with an elevated odds of testing positive for SARS-CoV-2 (OR=1.362, 95% CI=1.021 - 1.817, p-value 0.036; Table 4). However, when ApoA, HDL or BMI were controlled for, this effect was no longer significant (Table 4). In contrast to prior studies, we did not find any association between LDL levels and odds of testing positive for SARS-CoV-2 (OR = 0.995, 95% CI = 0.939 – 1.055, p-value = 0.872; Table 4) (9). Consistent with this, apolipoprotein B, the primary lipoprotein associated with plasma LDL, was not associated with any significant effect on odds of testing positive for SARS-CoV-2 (OR = 1.003, 95% CI = 0.947 – 1.063, p-value = 0.910; Table 4). Additionally, no significant effect of triglyceride levels on odds of testing positive for SARS-CoV-2 were found (OR = 1.026, 95% CI = 0.969 – 1.087, p-value = 0.375; Table 4).

**Table 4.**
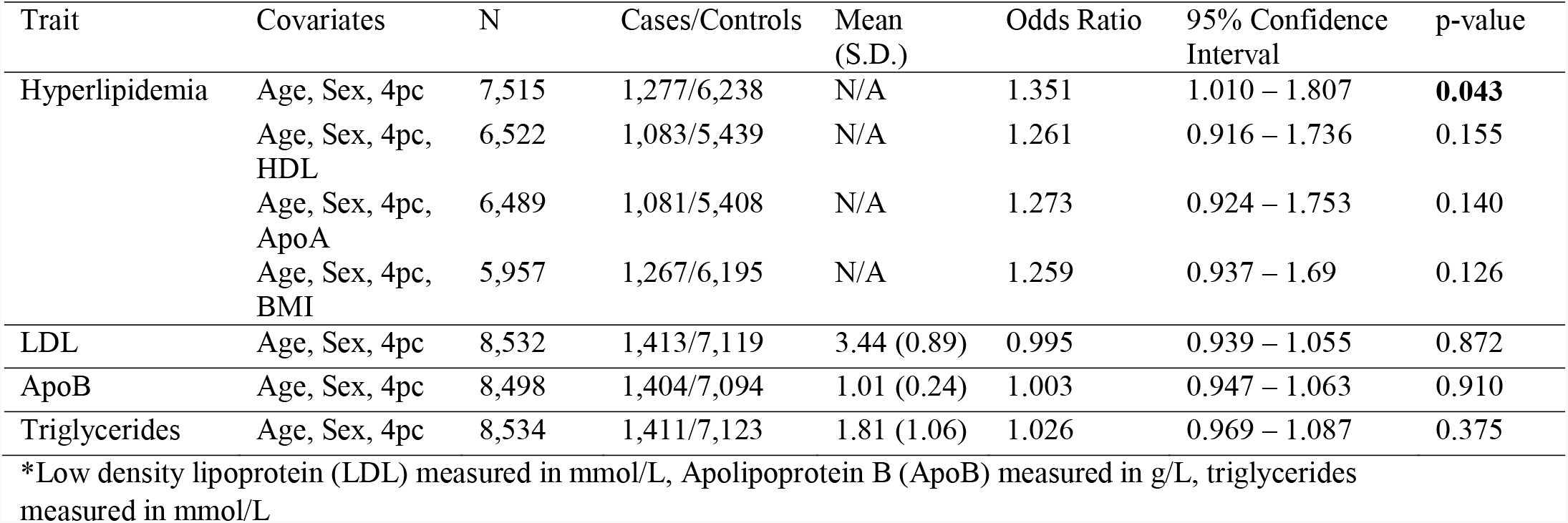
Effect of Hyperlipidemia, LDL, ApoB and Triglycerides.

### Alcohol/Red Wine Intake

Frequency of alcohol intake was associated with reduced odds of testing positive for SARS- CoV-2 (Figure 1). There appears to be a “dosage dependent effect” as the significant OR reduction is found only above 3-4 alcohol drink consumption per week (OR = 0.716, 95% CI = 0.578 - 0.997), p-value = 2.28×10^−3^, Table 5), with maximal impact seen with daily alcohol consumption (OR = 0.660, 95% CI = 0.530 – 0.821, p-value = 1.98 x 10^−4^; Table 5).

**Table 5.** Effect of Alcohol Usage Frequency.

| Alcohol Intake | Covariates | N | Cases/Control<br>s | Odds Ratio | 95% Confidence Interval | p-value |
| --- | --- | --- | --- | --- | --- | --- |
| 3 to 4x a week<br>vs. never | Age, Sex, 4pc | 2,774 | 456/2,318 | 0.716 | 0.578 – 0.997 | **$2.28 \times 10^{-3}$** |
|  | Age, Sex, 4pc,<br>HDL | 2,392 | 385/2,007 | 0.752 | 0.593 – 0.953 | **0.018** |
|  | Age, Sex, 4pc,<br>BMI | 2,752 | 452/2,300 | 0.746 | 0.601 – 0.927 | **$8.31 \times 10^{-3}$** |
| Daily vs never | Age, Sex, 4pc | 2,750 | 431/2,319 | 0.660 | 0.530 – 0.821 | **$1.98 \times 10^{-4}$** |
|  | Age, Sex, 4pc,<br>HDL | 2,363 | 367/1,996 | 0.733 | 0.574 – 0.936 | **0.013** |
|  | Age, Sex, 4pc,<br>BMI | 2,728 | 428/2,300 | 0.694 | 0.556 – 0.866 | **$1.25 \times 10^{-3}$** |

\*Alcohol intake assessed by questionnaire with multiple choice answers. Question was “About how often do you drink alcohol?” all answers compared to those who chose “never”

**Figure 1.**
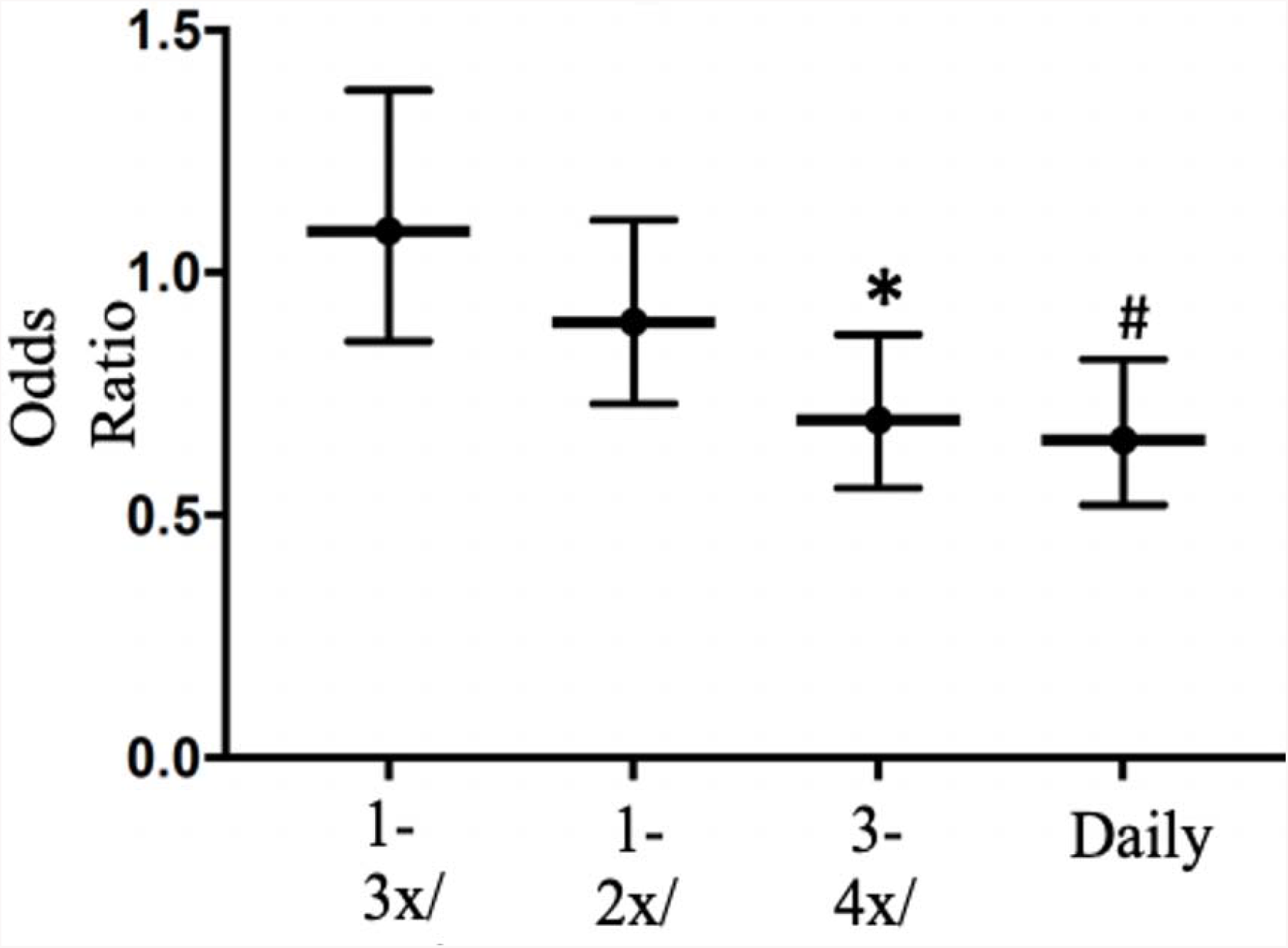
Dose dependent effect of alcohol consumption on SARS-CoV-2 positive testing risk. Alcohol intake assessed by questionnaire with multiple choice answers. Question was “About how often do you drink alcohol?” All answers compared to those who chose “never”. *Odds-ratio = 0.716, 95% CI = 0.58 - 0.89, p-value = 2.28×10^−3^ ^#^Odds-ratio = 0.660, 95% CI = 0.53 - 0.82, p-value = 1.98×10^−4^

Interestingly, this effect is diminished slightly after controlling for HDL (3-4x/week: OR = 0.752, 95% CI = 0.593 – 0.953, p = 0.0183; daily: OR = 0.733, 95% CI = 0.574 - 0.936, p-value = 0.0129; Table 5). Of the types of alcohol beverages recorded (Spirits, Fortified Wine, Red Wine, White Wine and Beer), only increased red wine intake was associated with reduced odds of testing positive for SARS-CoV-2 (OR = 0.883, 95% CI = 0.811 – 0.960, p-value = 3.75×10^−3^; Table 6). Again, this effect is diminished slightly after controlling for HDL (OR = 0.904, 95% CI = 0.825 – 0.991, p-value = 0.0314; Table 6), suggesting that HDL, at least in part, mediates the effect of alcohol intake on the odds of testing positive for SARS-CoV-2.

**Table 6.**
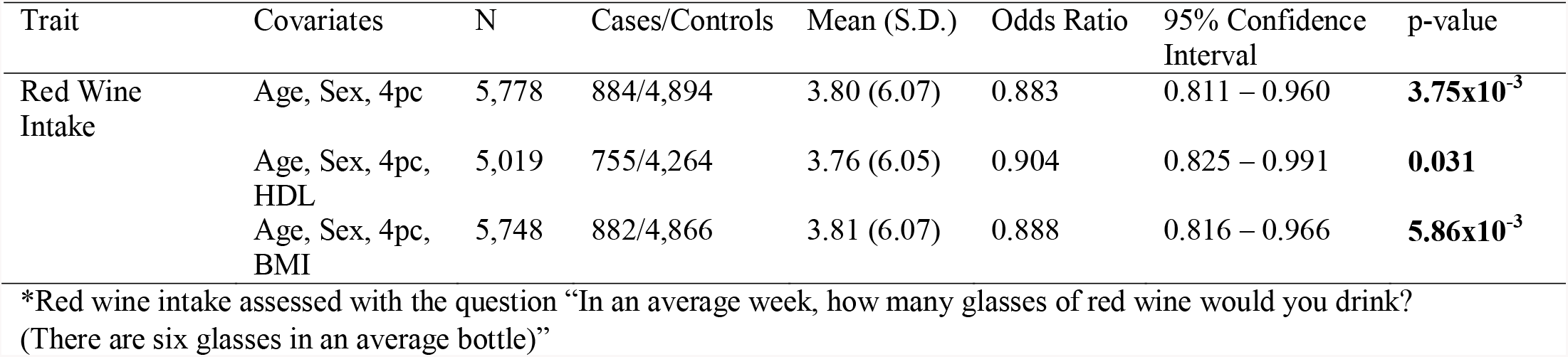
Effect of Red Wine Intake.

### Diabetes/HbA1c

The diagnosis of Type II diabetes (Phecode phe250.2) was associated with an elevated odds of testing positive for SARS-CoV-2 (OR =1.213, 95% CI =1.028 - 1.432, p-value = 0.0225; Table 7). Consistent with this, we found that HbA1c level was associated with a significant increase in odds of testing positive for SARS-CoV-2 (OR = 1.061, 95% CI = 1.005 – 1.121, p-value = 0.0332; Table 7). However, when ApoA, HDL or BMI were controlled for, the effects of both the type II diabetes diagnosis and HgA1c level were no longer significant (Table 7). Curiously, Type I diabetes (Phecode phe250.1) was not associated with an elevated odds of testing positive for SARS-CoV-2 (Table 7).

**Table 7.** Effect of Diabetes and HbA1c.

| Trait | Covariates | N | Cases/Controls | Mean (S.D.) | Odds Ratio | 95% Confidence Interval | p-value |
| --- | --- | --- | --- | --- | --- | --- | --- |
| Type II Diabetes | Age, Sex, 4pc | 8,948 | 1,500/7,448 | N/A | 1.213 | 1.028 – 1.432 | **0.023** |
|  | Age, Sex, 4pc, HDL | 7,770 | 1,283/6,487 | N/A | 1.131 | 0.943 – 1.356 | 0.184 |
|  | Age, Sex, 4pc, BMI | 8,882 | 1,488/7,394 | N/A | 1.126 | 0.947 – 1.34 | 0.180 |
| HbA1c* | Age, Sex, 4pc | 8,508 | 1,421/7,087 | 37.32 (8.43) | 1.061 | 1.005 – 1.121 | **0.033** |
|  | Age, Sex, 4pc, HDL | 7,398 | 1,217/6,181 | 37.39 (8.51) | 1.035 | 0.974 – 1.100 | 0.265 |
|  | Age, Sex, 4pc, BMI | 8,444 | 1,409/7,035 | 37.30 (8.41) | 1.03 | 0.972 – 1.09 | 0.317 |
| Type I Diabetes | Age, Sex, 4pc | 8,006 | 1,311/6,695 | N/A | 0.817 | 0.529 – 1.261 | 0.360 |

\*HbA1c measured in mmol/mol

## Discussion

Early studies of SARS-CoV-2 pandemic identified pre-existing cardiovascular disease and obesity as risk factors associated with acquiring SARS-CoV-2 (2-8). In addition prior studies found that hypolipidemia and declining HDL in the setting of acute Covid-19 illness was associated with disease severity (9, 10). Moreover, obesity was associated with higher prevalence of SARS-CoV-2 infection and Covid-19 disease severity (3, 7, 8). Here, we sought to validate these findings by examining the potential effects of baseline BMI, lipoproteins, their respective apolipoproteins and diabetes on the odds of acquiring SARS-CoV-2 in a cohort from the UK Biobank dataset. We confirmed that baseline hyperlipidemia was associated with the odds of testing positive for SARS-CoV-2, but this association was driven primarily by association of baseline lower HDL and ApoA levels with SARS-CoV-2 positivity, suggesting specifically that HDL may play a role in predisposing patients to SARS-CoV-2 infection. Moreover, consistent with an earlier study (7), we confirmed the association of high BMI with the odds of testing positive for SARS-CoV-2. Importantly, this effect was no longer significant when baseline HDL and ApoA levels were controlled for. We also found association of Type II (but not Type I) Diabetes and HbA1c levels with SARS-CoV-2 diagnosis. Again, when baseline ApoA or HDL levels were controlled for, the effects of both the Type II diabetes diagnosis and HbA1c level were no longer significant. Given that Type II diabetes is associated with reduced HDL levels, we hypothesize that the elevated odds associated with Type II diabetes is mediated in part through its effect on HDL levels. Taken together, our results suggest that HDL may be mediating part of the well known effect of BMI on SARS-CoV-2 /Covid-19 risk and plays a greater role in SARS-CoV-2 pathogenesis than previously appreciated.

Interestingly, we found that intake of red wine, but not other alcoholic beverages, was associated with reduced odds of testing positive for SARS-CoV-2 in a “dose-dependent” manner (at least 3 glasses per week). This is counterintuitive, given that prior research suggests there is a dose dependent relationship between alcohol intake and viral infections (17). Nevertheless, red wine, in particular, has been shown to have positive effects on endothelial function, glucose metabolism and HDL upregulation when consumed in moderate quantities (18). We speculate whether red wine intake and HDL may modulate inflammatory, endothelial and lipid characteristics in such a way to confer some degree of protection to SARS-CoV-2 infection.

Although the findings of this study persist when appropriate controls are applied, we acknowledge the inherent limitations of this association study, which is subject to sampling bias. Importantly, we do not know the context in which the SARS-CoV-2 testing was carried out, the HDL status at the time of testing, and the disease severity of each case. Participants tested in this study were primarily those who presented to a clinical care site with symptoms suggestive of SARS-CoV-2. Although this has the potential to marginally increase SARS-CoV-2 positive testing rate, it is unlikely to influence the association of HDL, BMI and alcohol consumption with SARS-CoV-2 positivity rate. Additionally, while majority of those tested were inpatient at the time of sampling, we acknowledge the potential confounding effects of subsequent expansion of testing into the community and to asymptomatic patients; however, we believe that such effects would tend to diminish any association with SARS-CoV-2 test positive rates.

Another limitation of an association study based on UK Biobank is that the baseline cardiometabolic data was collected several years prior to the SARS-CoV-2 pandemic. However, we do not believe that this did not have a substantial impact on our findings. While mean BMI has been reported to have increased around the world from 1976 to 2016, this increase has plateaued in recent years in high-income English-speaking countries, including the UK (22). Additionally, mean triglyceride, HDL and LDL levels tend to change only modestly or remain relatively stable in the population over time (23, 24). We believe that these modest changes over time would tend to diminish their associations with SARS-CoV-2 test positive rates, particularly if abnormal baseline levels were treated in the interim with medications. With regard to alcohol intake, the Institute of Alochol Studies based in the UK, reports that levels of drinking among adults in 2018 was comparable to drinking in 2010 (25).

Despite these limitations, we believe that our exploration of SARS-CoV-2 corroborates some of the earlier studies and provides valuable insight and guidance for future studies. One of the most compelling finding of this study is the association of lower baseline HDL levels with SARS-CoV-2 positivity, which corroborates an earlier study, which found association of declining HDL levels with Covid-19 disease severity (10). While causal inferences are beyond the scope of this study, given HDL’s established antioxidant, antithrombotic and anti-inflammatory characteristics, it seems plausible that HDL may play a protective role in preventing the establishment of SARS-CoV-2 infection (19). Alternatively, HDL is now known to transport a wide range of cargo other than cholesterol, such as microRNAs (miRNAs) (20). These non-canonical roles of HDL are consistent with the direct, nonselective antiviral effects of HDL (21). Our analysis opens up several avenues for further study, for example, to determine whether baseline HDL levels can identify high risk patients, to explore whether pharmacological modulation of HDL levels may confer protection against SARS-CoV-2 infection, or even to examine whether HDL particle confers direct protection against SARS-CoV-2 infection.

## Data Availability

All data used in this research was downloaded from the UK Biobank Resource under Application Number 49852. Copies of the data may be obtained from the UK Biobank Resource. The data access procedure is provided on their website. ( https://www.ukbiobank.ac.uk/wp-content/uploads/2012/09/Access-Procedures-2011-1.pdf )

## Acknowledgements

This research was conducted using the UK Biobank Resource under Application Number 49852. This work was supported by 5T32GM092237-10 to RS, NIGMS R01GM118557, NHLBI R01HL135129 to CCH, and NHLBI 1U01HL137181 to JP. The funders had no role in the design and conduct of the study; collection, management, analysis and interpretation of the data; preparation, review or approval of the manuscript; or decision to submit the manuscript for publication.

